# Barriers and facilitators to hepatitis C patient engagement: interview study with General Practitioners in England

**DOI:** 10.1101/2025.11.17.25340431

**Authors:** Avelie Stuart, Carina Hörst, Dolores Mullen, Catherine Lowndes, Ruth Simmons, Aneesha Noonan, Monica Desai

## Abstract

There are people living with a previously diagnosed chronic hepatitis C infection who have been lost to follow up, and people who may have undiagnosed hepatitis C. General Practitioners (GPs) could help to support these patients into care, via offering testing, referring to secondary care, and facilitating prescribing. We conducted semi-structured interviews with 14 GPs, most of whom work with local Operational Delivery Networks (ODNs) with funding from the NHS England Viral Hepatitis Programme as hepatitis C GP Champions. A thematic analysis with a multi-level model (system, provider-levels) and the COM-B (capability, opportunity, motivation: behaviour) framework were used to identify barriers and facilitators to patient identification and (re)engagement. System-level barriers included a lack of hepatitis C incentives for GPs, pressures facing primary care, challenges searching patient records, and a perception that better processes are needed to re-engage patients lost to follow up. GP Champions expressed challenges coordinating regional patient (re)engagement strategies, but also reported successful collaborations with secondary care, other services, and outreach/community partners. Provider-level barriers for all GPs included lack of knowledge of hepatitis C and difficulty managing patient conversations (capability); and lack of GP capacity and difficulty contacting patients (opportunity). Provider-level facilitators included that primary care is holistic and accessible to patients (opportunity), and GP Champions reported how their roles have granted workload capacity and the support of other GPs/healthcare professionals (opportunity) and increased their feelings of personal reward (motivation). Interviewees’ reflections on shared care and their suggestions for the future of hepatitis C care are also presented. This study demonstrates numerous benefits to having GP Champions employed to find and offer testing to patients, however system-level barriers are notably persistent. Interviewees said there is further need to address patients holistically, and joint efforts between services/sectors were suggested as means to overcome system and provider level barriers.

## Introduction

Following the introduction of highly-effective and well-tolerated Direct Acting Antivirals (DAAs, given orally, as part of a 8-12 week treatment course [1]), the WHO developed a Global Health Sector Strategy for the elimination of viral hepatitis by 2030 [2]. The UK Government has committed to meeting the elimination targets. In England there has been a decrease of 56.7% of people living with chronic hepatitis C since 2015, leaving an estimated 55,900 people who are unaware of their infection or have not been successfully treated or cleared the virus [3].

Primary care was not the initial focus of the elimination campaign in the UK. However, primary care providers (GPs) are the main point of healthcare access for many people in the UK. Moreover, GPs possess entire and continuous medical records, which means they can play a bigger role in identifying people with hepatitis C [4, 5]. Recent initiatives in England primary care settings include a digital search tool pilot that identifies people with historical records of HCV infection, or risk indicators [6]. In addition, a GP Champion initiative funded by the NHS England Elimination Programme is supporting around 50 GPs to take on a role in finding patients. In this context, it is important to understand the barriers and facilitators to identifying and (re)engaging people living with hepatitis C via primary care.

### Hepatitis C testing in primary care

GPs in the UK are recommended to offer testing to people at increased risk of hepatitis C, particularly migrants from medium- or high-prevalence countries and people who inject or have injected drugs [7]. However, GPs also have many competing priorities, and financial incentives are received for prioritising and achieving other targets in patient care. A Europe wide study found that GPs in the UK reported regularly offering testing for hepatitis C virus (HCV) to people who inject drugs, sex workers, and migrants [8]. Testing for HCV in patients with abnormal liver results was also fairly common. Yet, over half of UK GPs said they were rarely or never involved in the clinical management of hepatitis C, and specialists reported rarely referring patients back to GPs for management [8]. A separate UK study found that 27% of primary care offered new migrant patients HCV testing [9].

To boost testing rates, some targeted testing trials have been delivered in primary care and found to be cost-effective [10–12]. One study engaged primary care in searches for patients with hepatitis C risk factors in their patient record [13]. They found that GPs knew some of the risk factors but did not always offer tests based on these risk factors; that GPs did not always remember to regularly re-offer testing; and that hepatitis C was a lower priority for GPs than many other medical issues. Further, they found an unmet need to build rapport with patients as well as difficulties finding computer records. Some GPs saw other services as more suitable for hepatitis C care and reported that some patients seen in primary care are reluctant to engage or unable to engage with care. Similarly, in the U.S. a study with primary care providers found that they were aware of screening recommendations but other activities were prioritised [14].

In this study we aim to further explore the barriers and facilitators for GPs to offer testing to patients.

### Re-engagement of hepatitis C patients with historical diagnoses

In addition, we aim to understand the barriers and facilitators to re-engaging patients with historical diagnoses. Some people have a note of hepatitis C infection on their patient record but have not received treatment or cleared the virus; many of these were diagnosed at a time when treatments were less effective and less tolerable. Several exercises have been conducted with the aim of re-engaging patients [15–18]. For example, in England an exercise was carried out by secondary care (the NHS Operational Delivery Networks (ODNs)) from 2018 onwards using historical laboratory records [15, 19]. The qualitative evaluation of this exercise [20] found that ODNs had difficulty making contact with many patients and lacked information that could have been provided by GPs (although they acknowledged the pressures faced by GPs); further, they believed that GPs might be more suited to support patients who were originally diagnosed in primary care and/or who are lost to follow-up.

Re-engaging historically diagnosed patients may require understanding and addressing various factors that impede their access to healthcare, and they may have additional needs [21]. For example, people living with homelessness or people from migrant communities, who have been excluded from care or are not well served by conventional care, may need additional or tailored support [20]. Primary care providers may be positioned to support these patients. Previous international research has found that primary care providers can motivate patients to consider the broader outcomes from having treatment [22–25], can help address myths of ineligibility for treatment (e.g. due to ongoing drug use), and can build trust with patients to reduce internalised stigma [22, 23].

### Hepatitis C treatment in primary care

Some countries have also attempted to decentralise hepatitis C treatment to primary care [e.g. 26, 27, 28], since DAAs require less specialist skills to administer. Some patients may find primary care more accessible as the first point of contact [29]. A study in Scotland [28] found barriers to decentralising care, including a need for patients to be assessed for liver fibrosis (which is not part of standard GP contracting), lack of GP capacity, and that hepatitis C was viewed by GPs as a specialty treatment. Facilitators included the relative simplicity of DAA treatment, good GP/patient relationships, and holistic care approaches. A synthesis of qualitative literature among identified priority populations for hepatitis C (people who inject drugs, men who have sex with men, indigenous people) in high income countries [22] found that the main provider (including primary, secondary and other sectors) barriers to delivering hepatitis C treatment were lack of resources, lack of knowledge of, and confidence in, delivering treatment, as well as reticence to treat ‘difficult’ patients. The facilitators they identified were the simplicity of treatment with DAAs, co-location of hepatitis C care with other health services, training for boosting provider confidence, and how their professional identity as a doctor boosted motivation to provide treatment.

### The current study

This study employs expert interviews [30] with GPs experienced in hepatitis C. The aim is to understand barriers and facilitators to identifying, (re)engaging, and potentially treating patients living with hepatitis C via primary care in England. We use a socio-ecological model to understand barriers and facilitators at system and provider levels [22, 25], further employing the behavioural COM-B framework (Capability, Opportunity, Motivation – Behaviour [27, 28, 31]), to classify barriers and facilitators at the provider-behaviour level.

## Method

### Design and setting

Semi-structured expert interviews with GPs were chosen as they allow an in-depth exploration of barriers and facilitators with professionals experienced in the subject [30]. An NHS England-commissioned hepatitis C GP Champion programme is operating nationally. There are currently approximately 50 GP Champions in post. GP Champions are paid to work with the ODNs to conduct hepatitis C training, patient record searches, and to hold events to find and engage patients across Primary Care Networks (PCNs). They have been being recruited by ODNs since 2022 and currently 18 out of the 25 ODNs have at least one GP Champion. The aim was to achieve a sample size of 15-20 GPs [32]. We adhered to the standards for reporting qualitative research (SRQR) in conducting the research and writing the manuscript [33].

### Materials

The interview topic guide covered three broad areas. Section 1 included questions about participants’ roles as GPs, their role in hepatitis C care, and their knowledge of the availability of hepatitis C training resources. Sections 2 and 3 were developed using the COM-B model (Figure 1) [31], with section 2 focusing on barriers and facilitators to the (re)engagement of historically-diagnosed hepatitis C patients, and Section 3 on barriers and facilitators to routine hepatitis C care (e.g. testing patients). The COM-B model was used in the topic guide to frame questions about the drivers of GP behaviour in terms of Capability (including psychological and physical skills, knowledge), Opportunity (including physical environment and resources, social influences, relationships and social norms), and Motivation (reflective motivation - including planning, thinking, and evaluation, and automatic motivation - involving emotional and intuitive reactions and habits). We also checked our topic guide against the Theoretical Domains Framework (TDF) [34], which breaks down the COM-B into further specific domains, to ensure adequate coverage of potentially relevant behavioural drivers.

**Fig 1.**
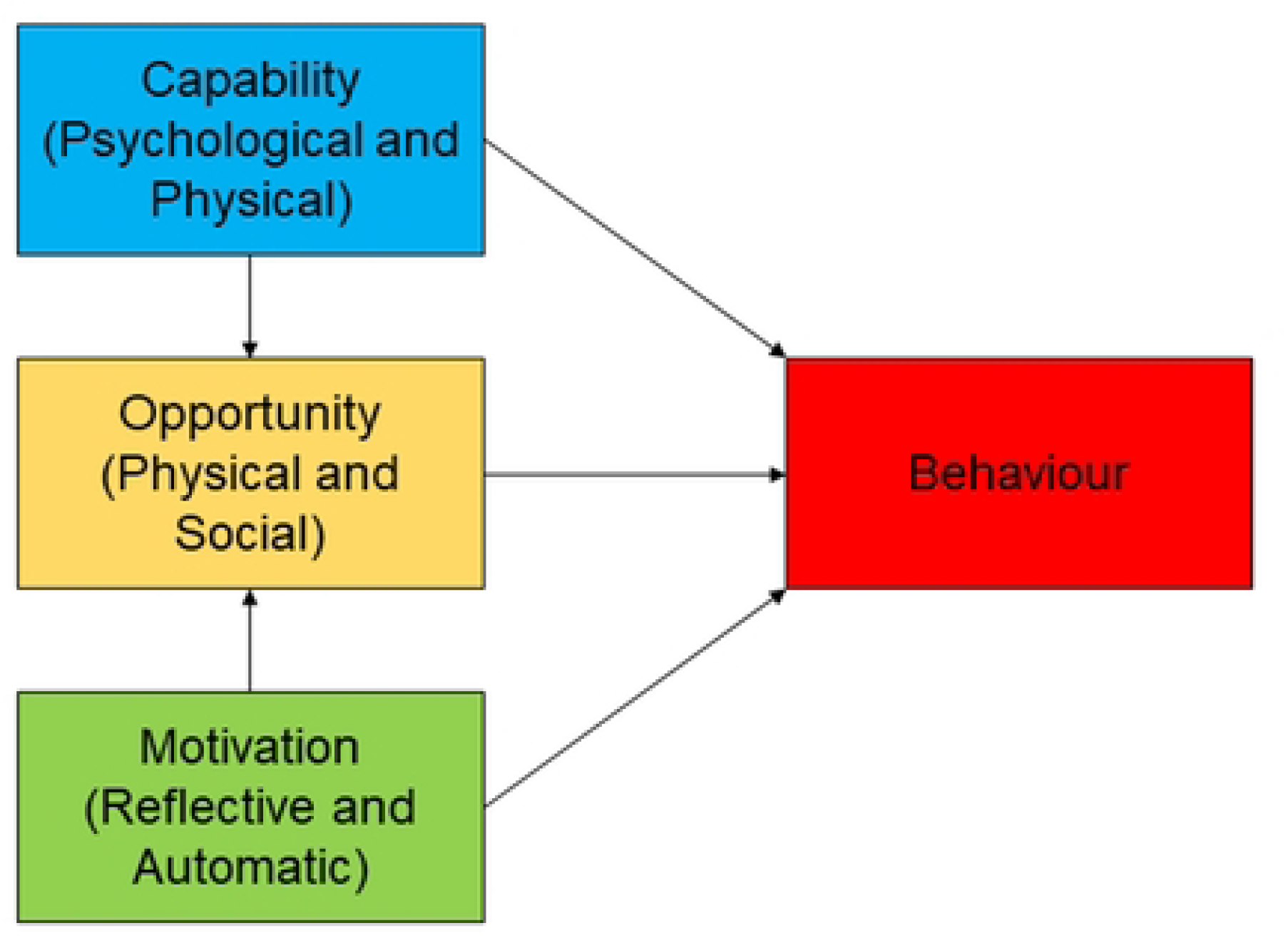
The COM-B model [31], depicting how capability, opportunity and motivation contribute to behaviour.

Example interview questions are included in Table 1 (see S1 Appendix for topic guide).

**Table 1.**
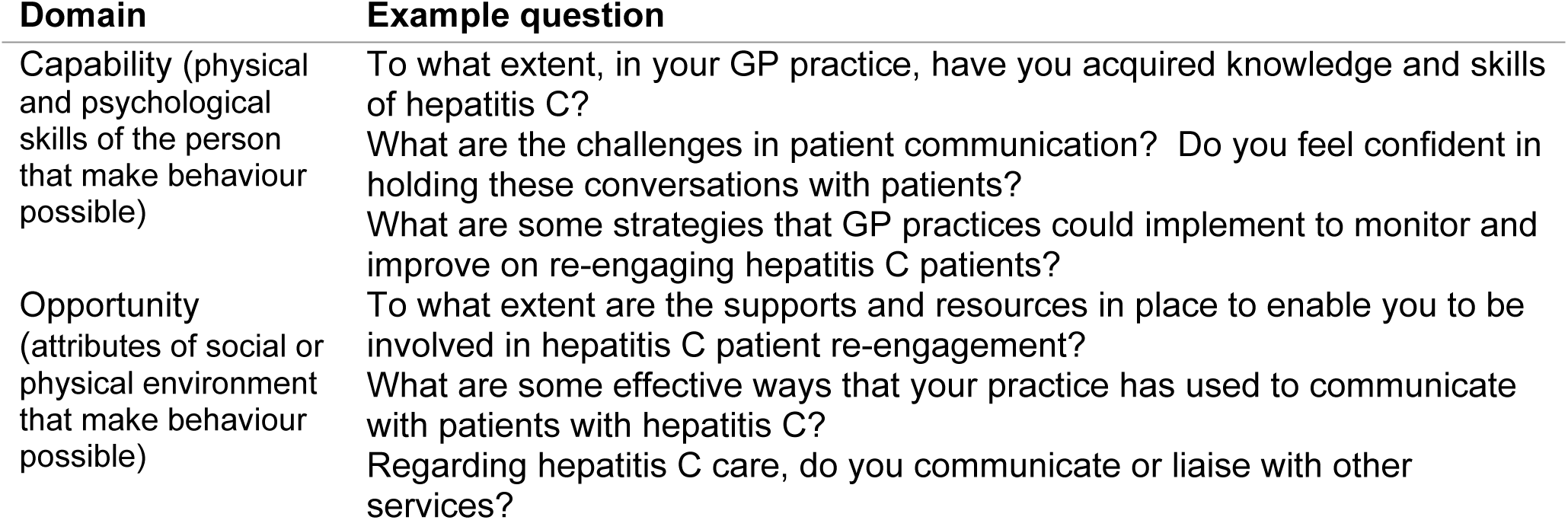
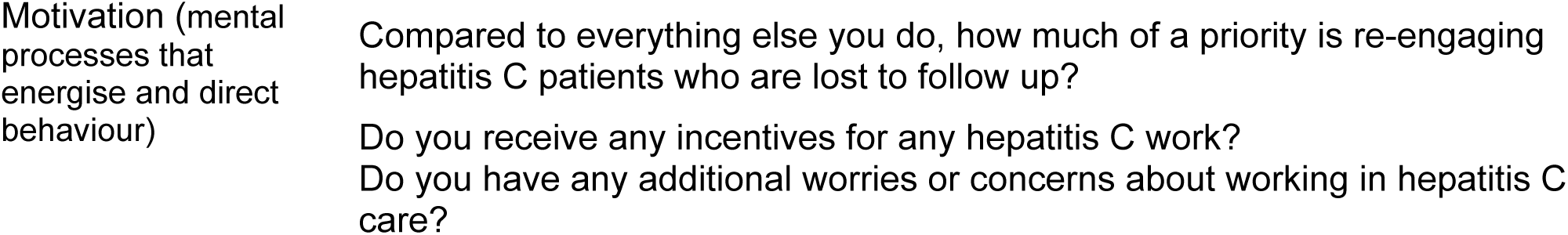
Example topic guide questions per COM-B domain.

### Procedure

We used convenience and snowball approaches for recruitment. GPs on the NHS hepatitis C GP Champion email list were first emailed 07^th^ June 2024, and we advertised the study in an organisational newsletter for GPs. ODNs who had previously indicated they were working with GPs [20] were also asked to distribute the advert. All participants were asked to distribute the study advert amongst their networks. Data collection ended 17^th^ September 2024. The interviews were conducted via Microsoft Teams, lasted between 30 minutes to 1 hour 10 minutes (average 44 minutes), and were recorded for transcription purposes only.

### Data analysis

Analysis was undertaken using NVivo 14, involving thematic analysis [35] with combined deductive and inductive processes (see Figure 2). The deductive analysis structure is adapted from previous hepatitis C research [22, 25] and the COM-B model [31]. Then themes were inductively generated as either barriers, facilitators or mixed themes. A barrier was defined as a factor that hinders/obstructs, a facilitator as a factor that promotes/enables/supports, and mixed themes act as both barriers and facilitators. System level themes are where the interviewee attributes responsibility to systems rather than provider agency/behaviour; and likewise, provider-level themes are those that interviewees attributed to provider agency rather than systems.

**Fig 2.**
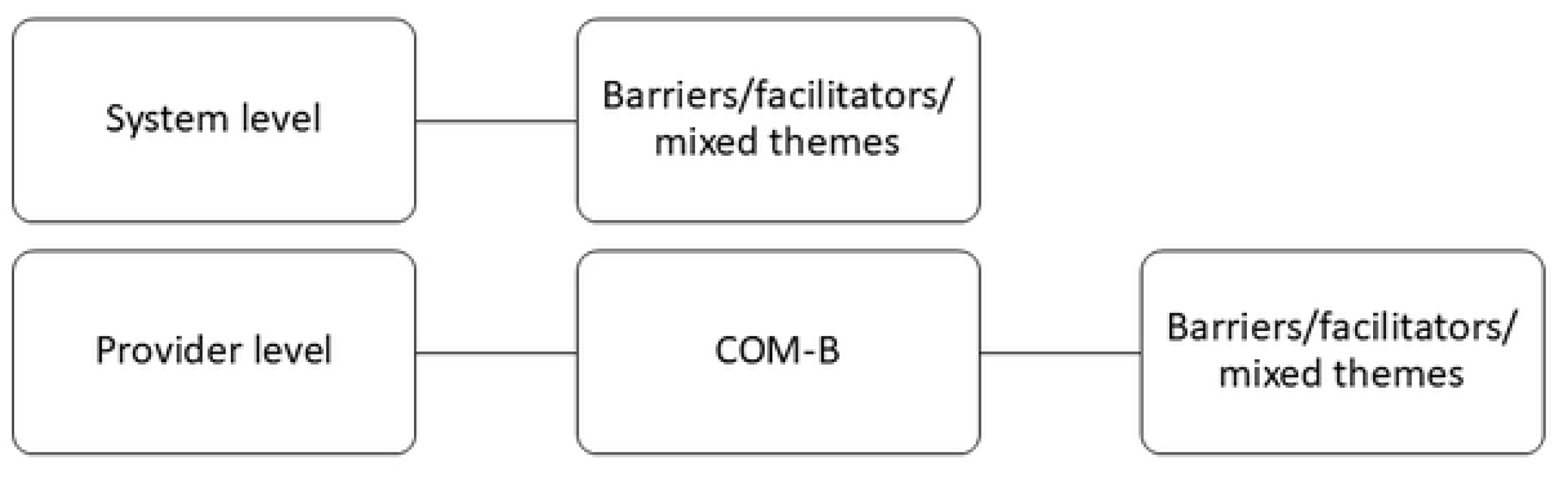
Deductive and inductive coding structure.

The analysis involved the following steps: 1) transcripts were re-read for familiarisation, 2) codes were created and assigned deductively, 3) codes were reviewed for meaning and inductively grouped into themes, 4) themes were reviewed by CH and revised by AS until agreement was reached, and finally, 5) the manuscript was written with example quotes provided. We also coded barriers/facilitators that the interviewees attributed to patients’ experiences, and for thoroughness include these in the S2 Appendix. Provider perspectives about patients may be useful, for example, to inform provider training - but are not equivalent to hearing directly from patients. In addition, the topics of ‘shared care’ and ‘primary care prescribing treatment’ arose in a small number of interviews and so we produced a subsample analysis of these themes. Finally, we identified themes from interviewees’ suggestions for future practice.

### Patient and public involvement

This study does not involve patient or public involvement; however, is part of a series of studies on HCV patient (re)engagement, which has involved lived experience representatives from the Hepatitis C Trust. The topic guide was piloted with four public health professionals, enabling improvements to the relevance and flow of questions.

### Ethics

The study was approved by the UKHSA Research Ethics and Governance Group (ID: 575). Participants were informed about the study and provided electronic written consent.

## Results

### Participants

We interviewed 14 GPs, 10 based in London, three in north England, and one in the south-west of England. Their experience as practising GPs ranged from 1 to more than 30 years. All but two participants were hepatitis C GP Champions. All Champions had regular GP roles in addition to their Champion role.

### System level barriers and facilitators

Table 2 contains the main barriers and facilitators identified at a system level. Themes were chosen for inclusion in the main manuscript based on either frequency or expressed importance for hepatitis C care.

**Table 2.** Summary of system level themes (further themes are in the S2 Appendix).

| All GPs or specific<br>to GP Champions | Theme | Barrier,<br>Mixed,<br>Facilitator | Number of<br>Participants | Related to patient<br>identification, re-engagement or<br>both? |
| --- | --- | --- | --- | --- |
| All GPs | No financial incentives for hepatitis C in primary care | Barrier | 7 | Both |
|  | Pressures facing primary care | Barrier | 11 | Both |
|  | Previous treatments were associated with disengagement from care | Barrier | 5 | Reengagement |
|  | Lack of standard procedure for re-engaging patients in primary care | Barrier | 8 | Reengagement |
|  | Patient search tools and hepatitis C codes in GP practice systems | Mixed | 14 | Both |
| GP champions | Challenges collaborating with primary care networks and regions | Barrier | 8 | Both |
|  | Uncertain strategy for contacting large numbers of patients for testing | Barrier | 9 | Identification |
|  | Collaborating with the Operational Delivery Networks (ODNs) | Mixed | 10 | Both |
|  | Collaborating with drug and alcohol services | Mixed | 3 | Both |
|  | Collaborating with outreach services/charities/partners | Facilitator | 5 | Both |

### System-level themes impacting all GPs

#### No financial incentives for hepatitis C in primary care ***and*** pressures facing primary care *(barriers)*

Interviewees emphasised the challenges facing GPs who have many competing priorities and are increasingly being asked to manage further chronic health conditions. These are well-known substantial barriers to practice, but not specific to identifying and (re)engaging patients living with hepatitis C.

#### Previous treatments were associated with disengagement from care (barrier)

Interviewees talked about a legacy issue that complicates patient re-engagement due to the relatively ineffective and poorly tolerated interferon treatment. Since DAAs became available, interviewees said that not all GPs have tried to re-engage these patients due to them needing additional support that is not usually provided by primary care (e.g. longer, flexible appointments):

> “we were still thinking about interferon treatment which had a very bad reputation. It was very difficult to use and then when Hep C treatment did come available, it was so expensive that we didn’t even have the opportunity to refer our patients. … So those immediate patients were picked up and treated, but the patients that might have needed more, more adaptability to clinic times and flexibility weren’t really treated…” (Participant 1)

#### Lack of standard procedure for re-engaging patients in primary care (barrier)

Interviewees said that GPs aim to be accessible to their patients and bear final responsibility for them, whereas secondary care can discharge patients. However, they also said that for hepatitis C typically they do not follow up patients after referral to secondary care which can result in patients being lost to follow up. In addition, as GP surgeries have a fixed catchment area and set number of sessions per week this limits their ability to follow up patients - especially when patients move away. This combination of factors led interviewees to say there were a lack of clear responsibilities/guidelines for GPs on re-engaging patients who have been lost to follow up.

> “I think there’s a sense that’s the responsibility that is on you and for how often do you chase people up or how often do you engage them? So I think that’s a very grey area.” (Participant 12)

#### Patient search tools and hepatitis C codes in GP practice systems (mixed)

Interviewees discussed electronic patient record search tools for finding patients with hepatitis C lost to follow up, or with risk factors for hepatitis C. Some interviewees said that the searches return a low number of patients with confirmed hepatitis C, whilst also returning a high number of patients with risk factors who have not been tested. Interviewees reflected on the infeasibility of tasking every GP in England to search for such patients.

> “[The PSI^1^ tool is] not a perfect search [tool] because it does pull in a number of risk factors and what that means is that in real terms you get you can get depending on where you work very, very, very high numbers and actually. You know, a lot of those patients. Won’t really need a test.…” (Participant 10)

Underpinning these issues were technology limitations (with electronic records and search tools) and the need for updates to the medical nomenclature for hepatitis C that the searches rely on. GP Champions were concerned about establishing a strategy for arranging these searches regionally, to alleviate standard GPs from having to conduct this work. GP Champion specific themes are expanded on in the next section.

### GP Champion-specific system level barriers and facilitators

#### Challenges collaborating with primary care networks and regions (barrier)

Some GP Champions had started planning regional patient search strategies, in collaboration with other primary care providers. They reported it as a lengthy process to create data sharing agreements and undertake governance reviews. One interviewee remarked that GPs are very protective of their patients’ privacy, need persuading that it would not be extra work for them, and that each practice will have their own views on the best way to approach this task.

> “[it] took a lot of time to get the data protection policies in place, data sharing policies in place going through the governance, clinical governance, Caldicott Guardians. But you know, so that did take a lot of meetings and a lot of documents, ultimately with patient care at the heart of it but you know, GP practices are very concerned with who has their patient data and how it gets shared and who contacts their patients.” (Participant 14)

#### Uncertain strategy for contacting large numbers of patients for testing (barrier)

The prospect of working across regions also caused GP Champions to reflect on the complexity of developing an appropriate patient communication strategy. Text messages were identified as the most feasible method. However, they were concerned about patient’s trust of and reactions to text messages (including feeling stigmatised), and whether to specify why people were contacted. Another concern was that this may not be an effective strategy.

> “we’ve been thinking about sending kind of mass text messages out, but there’ve been some pushback against that being people, patients not liking to receive them. And the wording around those and people sharing telephones. And is that text messages going to the right person? But if you make it too vague, nobody’s going to reply. But if you look at too specific and you’re at risk of breaking confidentiality.” (Participant 12)

Interviewees reported several ways they were considering reducing risks, including multiple follow up messages, testing the wording with patients, or softening the wording. Others were considering implementing opt-out or intake testing instead.

#### Collaborating with the Operational Delivery Networks (ODNs) (mixed)

Interviewees discussed the benefits of working with secondary care providers (ODNs), with shared responsibility for patient care and data sharing in place. Some ODNs were said to have been able to use their funding to incentivise GPs to refer patients, enabling GPs to create capacity in their workload. However, some interviewees encountered delays in setting up a partnership with the ODN due to capacity/resource limitations, and others expressed a concern that collaborating with ODNs would increase GPs workload.

> “it’s careful what you wish for isn’t it? But there it’s [the ODN] such a fantastic structure and it does, it’s very responsive. So in some ways if people did know about it, I wonder whether there’ll be more traffic because anything that makes a GPs job easier I think would be…” (Participant 12)

#### Collaborating with drug and alcohol services (mixed)

Locally commissioned drug and alcohol services manage hepatitis C care separately from primary care. This was said to reduce the demand on primary care but also result in GPs missing out on information on their patients status.

> “I have noticed a huge increase in [drug and alcohol services’] ability to manage positive cases over the last couple of years so they would manage that a positive Hep C RNA result and referrals treatment without any involvement of the GP which is the way it should be. So yeah, they tend to manage their Hepatitis C patients independently of us.” (Participant 3)

However, some interviewees reported good collaboration and communication with drug and alcohol services, where they had helped connect staff from each service or facilitated shared testing events in their locality.

#### Collaborating with outreach services/charities/partners (facilitator)

The most frequent facilitator at system-level was where interviewees had collaborated with outreach services, charities, and community groups to run joint testing events, such as a homeless health bus service, and ‘liver health’ events held in higher risk/prevalence communities. These events were said to have helped identify people living with hepatitis C.

> “I’ve linked colleagues from the Hep C Trust with our local charities, voluntary sector. In my boroughs, so in [areas] for the populations and ethnicities that we know are higher risk … we have a big deportation housing, in [Region] so we …they’re doing a lot of work out in the community” (Participant 14)

### Provider level barriers and facilitators

In this section we examine provider-level themes (listed in Table 3). We distinguish between themes specific to the GP champion role and those related to all GPs.

**Table 3.**
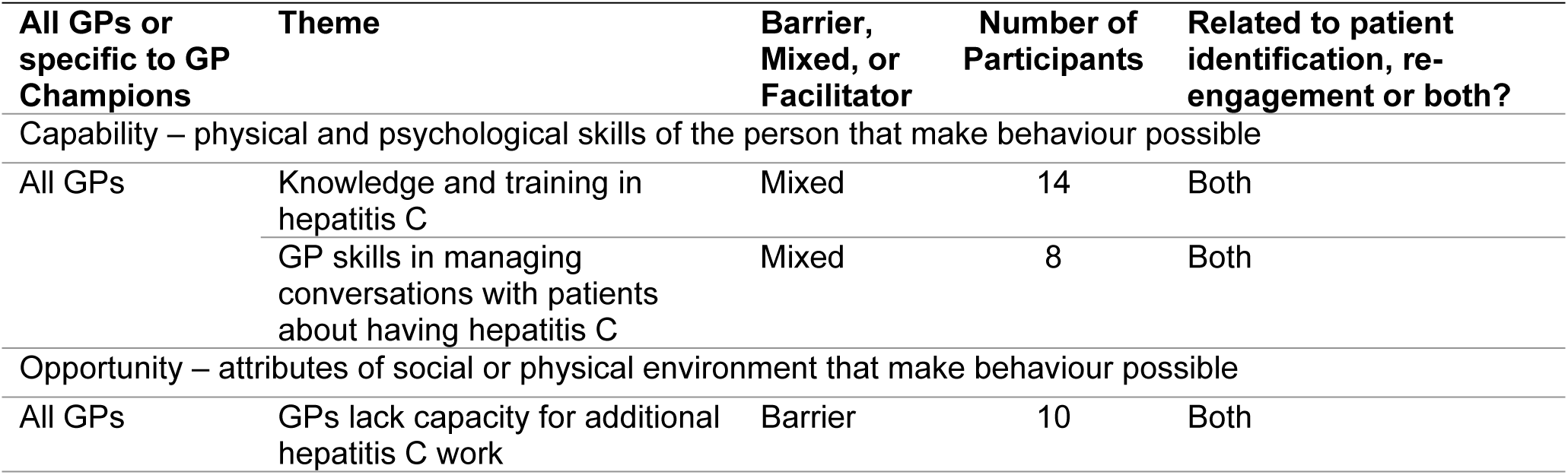
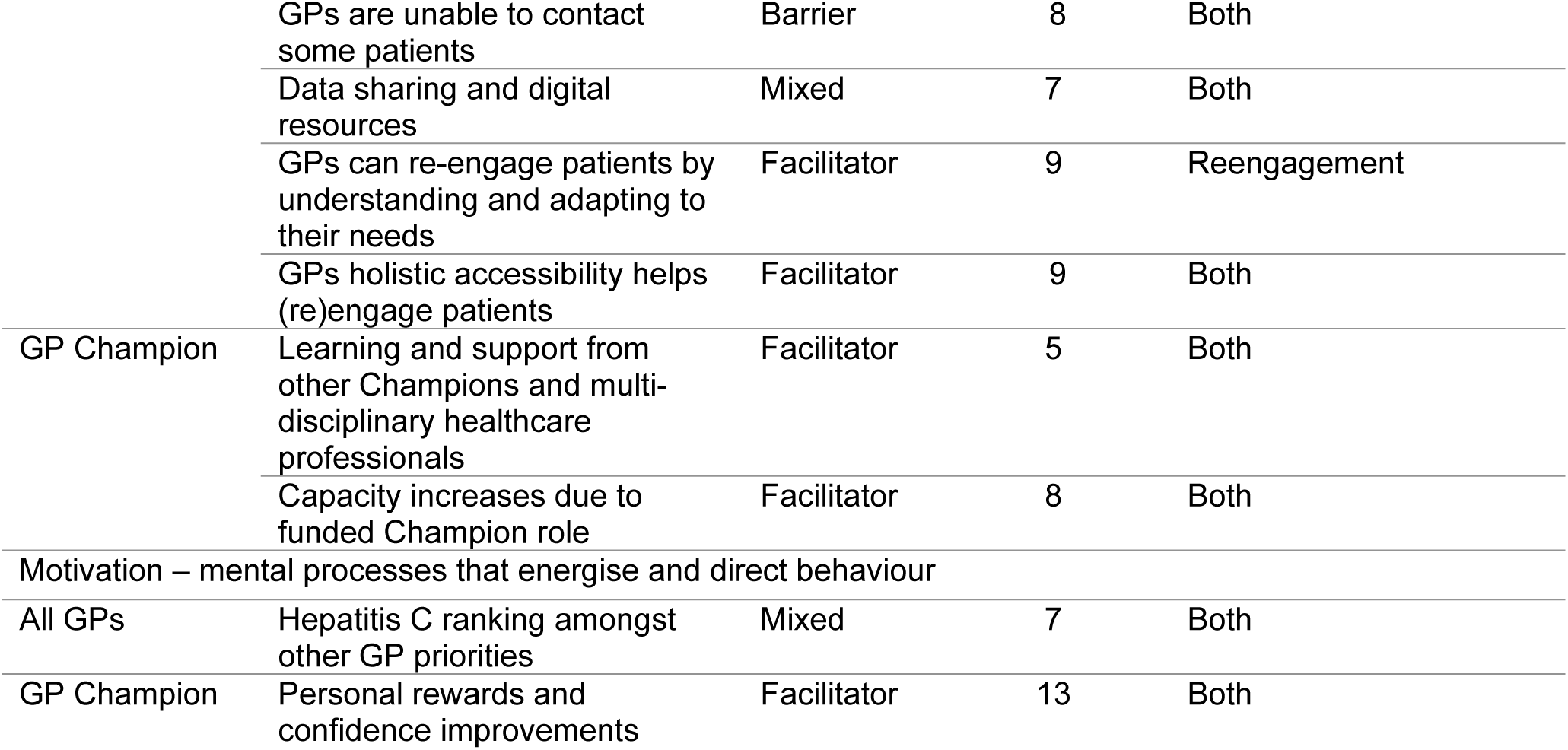
Provider-level barrier and facilitator themes, classified within the COM-B model (see S2 Appendix for full list of themes).

### Capability

#### Knowledge and training in hepatitis C (mixed)

Interviewees placed importance on the knowledge of GPs, nurses and other practice staff. Knowledge included when to test, how to access hepatitis C test records, and knowledge of the ODNs and patient referral procedures. Interviewees said concerns included GPs not knowing to test for hepatitis C when a patient has abnormal liver function test, and lack of knowledge that there is curative treatment.

> “I think a lot of the GPs then didn’t realise how Hep C treatment has become…. So, we have all these patients that we know in primary care are positive. However, we haven’t really referred them on or done anything about them because we’re, GPs, are still in that mindset.” (Participant 1)

However, some interviewees said they felt that knowledge/awareness amongst GPs is now adequate.

#### GP skills in managing conversations with patients about hepatitis C (mixed)

Some interviewees said that although personal conversations are part of being a GP they can still find it difficult to find the right time or approach to the subject, particularly as hepatitis C is a stigmatising condition.

“We have all these patients and it’s really really that are probably are eligible for Hep C tests, but we don’t do enough risk assessments in primary care, and that’s because it’s a really difficult subject to sometimes bring up.” (Participant 1)

GPs felt that with newer patients the conversation is easier for the GP as they do not have to address patients’ knowledge of previous treatment regimes.

### Opportunity

#### GPs lack capacity for additional hepatitis C work (barrier)

Linked to the pressures faced by GPs was a barrier in workload capacity. Lack of additional capacity was said to prevent GPs from being ‘proactive’ in identifying patients with hepatitis C or following up patients lost to follow up.

> “but the only problem is because GPs are if they’re overwhelmed, you know they can’t be proactive because they’re dealing with, you know, the clamour for appointments” (Participant 2)

#### GPs are unable to contact some patients (barrier)

Interviewees reported that identification and (re)engagement of patients is harder for GPs when patients change contact details or leave the area or country without notifying their GP.

> “…A lot of the time people have moved on … and they’re abroad and they can’t be contacted.” (Participant 3)

#### Data sharing and digital resources (mixed)

Interviewees reported a range of data sharing and digital resources that are available to them. For example, public health surveillance data can enable GPs to find out whether their patients have received care elsewhere. ‘Pop ups’ on patient record systems, automated blood test panels (so that GPs do not need to manually select the relevant blood tests), automated referrals and patient notification systems were all said to be useful, although in need of improvement. Additionally, specially developed hepatitis C websites, or a main practice website, were useful for keeping patients and GPs informed. However, some interviewees expressed concerns, for example, that these tools might cause ‘alert fatigue’, or prevent GPs from gaining knowledge if the processes are automated.

> “I think [the digital standard blood panel] is good because it increases testing but maybe that doesn’t help with awareness because people don’t know what they’re screening for.” (Participant 12)

#### GPs can re-engage patients by understanding and adapting to their needs (facilitator)

To re-engage patients, interviewees felt that it is necessary to spend time understanding the patient first and why they disengaged from care. Once doing so, they found they were better able to re-engage patients by offering suitable communication methods and flexible appointments based on their needs.

> “it’s having a flexible approach to bringing people in and making appointments is important. “(Participant 12)

#### GPs holistic accessibility helps (re)engage patients (facilitator)

Interviewees discussed how GPs, as the first point of contact for patients, are uniquely useful for patient (re)engagement because they have repeat opportunities that enable trust-building and are more locally accessible than secondary care.

> “we might have more of a relationship with people, there may be another reason that come here, they might come for medication, reviews or because they’ve got a sore throat and if you can tie it into that bring it up for a different reason while their there, then we have that advantage of the relationship and continuity.” (Participant 3)

#### GP Champions: Learning and support from multi-disciplinary healthcare professionals (facilitator)

GP Champions cited the support and networking benefits of meeting with and learning from other Champions, secondary care specialists, and multi-disciplinary healthcare professionals.

> “we have training like a training day once every six months? …it’s really useful. I think the most useful part is hearing is kind of the shared learning called case studies from other areas which is useful. And it’s also nice to be in a room full of other GPs who are interested in the same thing.” (Participant 12)

#### GP Champions: Capacity increases due to funded Champion role (facilitator)

Another facilitator reported by GP Champions is that their funded roles have granted them opportunities to prioritise hepatitis C. For example, enabling them the time to join with outreach services to run testing events; to communicate and strengthen relationships with local services and ODNs; to improve their own awareness; to work more flexibly; and to proactively contact patients.

> “I guess probably for me it’s [hepatitis C] relatively high up on my priority list, but that again, that is probably because I’m given the time and space to think about it and afforded that.” (Participant 3)

### Motivation

#### Hepatitis C ranking amongst other GP priorities (mixed)

Interviewees commented that most GPs have hepatitis C low on their list of competing priorities. Interviewees presented scenarios where other GPs might not be motivated to prioritise hepatitis C – such as not reading the letters informing them that their patient has not attended a hospital appointment or not prioritising a discussion of hepatitis C in a 10-minute appointment with a patient.

> “I don’t think anybody’s gonna do much chasing unless there’s a driver for it. …depending on how important the condition is, of course or how important the GP thinks the condition is, they might or might not chase them, but you know, if the people vanish, then they’ll probably shrug their shoulders.” (Participant 14)

#### GP Champions: Personal rewards and confidence improvements (facilitator)

Most of the Champions said their involvement in hepatitis C care is personally and professionally rewarding, especially as GPs do not often have opportunities to be directly involved in curative or preventative care; the Champion role reportedly facilitated these feelings of personal reward.

> “Yeah, it’s really satisfying. And I’ve learnt a lot about hepatitis C. Which again, yeah, from a, I guess a clinical, personal perspective. Very interesting. So I would say those are the main benefits and it’s just it’s not, it’s just again it’s variety in your working week.” (Participant 10)

### Prescribing in primary care or adopting shared care

Several interviewees discussed sharing care between primary care and secondary/other services, and/or treating individuals in primary care. As it was only discussed briefly, we produced a sub-sample inductive analysis.

The barriers identified to shared care or prescribing in primary care related to the additional administrative time and patient communication skills GPs would need to manage prescriptions and monitor patient adherence to treatment. Some perceived benefits, however, were that patients might prefer receiving treatment from primary care.

> “they might not be seeing secondary care, but sometimes they are coming into us and seeing primary care for various different reasons. And because of very different reasons may be chaotic lifestyle or it actually some patients recently that we’ve found by going through the notes, it’s just like, ‘oh yeah, I saw someone about it 15 years ago, and yeah, it’s not really a big thing and I don’t want to interferon treatment’ and they won’t go to secondary care, but they’re happy to keep seeing us in primary care” (Participant 1)

The ability for GPs to prescribe DAAs was seen as potentially enabling quicker linkage to treatment and a reduction in the people being lost through the referral process. The unique role of a GP (accessibility, holistic approach to patient needs) was seen as an advantage, especially if combined with other disciplinary support (e.g. pharmacy, housing support).

Some interviewees reflected on how they gained prescribing rights via the ODN. They stated this was a complicated and lengthy process, but positive once achieved. The DAA treatment being straightforward (tablet form, short course) was also said to enable GPs to prescribe DAAs. However, there were still concerns about the lack of provision of time and resources for GPs.

> “I work really closely with secondary care obviously in the ODN and everything like that. And I’m it’s taken a while, but I’m definitely part of their team now, even though that was a bit of a process…. I worry that in the transition over for primary care to have a greater role. Well, those two things [time and resource] might be. Not provided for enough.” (Participant 8)

### Interviewees’ suggestions for future hepatitis C care involving primary care

This final section summarises suggestions made by the GP interviewees for future hepatitis C care in primary care (see S2 appendix for full list of suggestions).

***Incentivising hepatitis C*** was suggested to enable hepatitis C testing to become embedded in routine practice, and to enable following up patients.

> “GPs are incentivised for all their long-term conditions and I know hepatitis C is not a long term condition because it’s curable, but I think if we do want to achieve micro eliminations and things, we need to incentivise the GPs practices to do the work and to do the testing.… even the fact that there would be a financial incentive, it just puts it back on them, you know, puts it back on the agenda with the other conditions like COPD, diabetes, asthma. That kind of thing, yeah” (Participant 5)

Additional ways of ***standardising regular opportunistic testing*** were emphasised by interviewees - if patients are regularly attending primary care for other purposes, they should be re-offered testing/treatment.

> “We do need to think of like systematic ways of embedding it. So, it becomes like routine. So, you know like when we test for abnormal LFTs [lateral flow tests], Hep C just in there rather than us having to actively think do I need to add in a Hep C test just automatic in there wherever we move away from risk assessments in high prevalence areas to just automatically testing things like that. Then maintains the focus, but in a subdued way,” (Participant 1)

Primary care was discussed as the main avenue for re-engaging patients who are not being offered tests elsewhere, such as drug and alcohol services. Interviewees said that GP patient record systems need changes to support such aims - for example, reminders and pop-ups to appear on patient records when the patient attends for another appointment. Others suggested it would be better to implement checks with all new patients on registration.

Interviewees discussed more generally the potential benefits and ***need for GP Champions who can support other GPs*** in identifying and (re)engaging patients or alleviating the GPs’ workload.

> “We’re only going to be finding like each individual GP, practise is only going to be finding, say like two or three patients maybe. So I still think that a lot of this needs to be done, not at a practise level, but at a systems level.” (Participant 8)

Suggested activities included running regional searches across practices and accessing other practices’ patient notes; meeting with Primary Care Networks (PCNs)/utilising PCN data managers and liaising between GPs and ODNs to create of data sharing agreements and honorary contracts; and giving GPs information and training so that they feel equipped to speak to their patients.

Suggestions were made for ***training GPs and other practice staff***, such as embedding hepatitis C into medical/GP training early, and production of small ‘bite size’ learning resources for time-poor professionals. Some interviewees stated that GPs working in areas of high need should have hepatitis C included as part of ***health inclusion principles***, due to the prevalence of hepatitis C in people living with homelessness, as well as migrants.

“I think potentially for GPs in the deprived area, it has to be a bit of a focus and has to fit in around kind of inclusion health principles, so inclusion health is the sort of care of people who have who have poor health outcomes from certain groups. So, people who experience in homelessness, asylum seekers, people learn[ing] disabilities, people in the prison service.” (Participant 3)

A ***joined-up approach*** with secondary care and charity sectors, and with other blood-borne virus elimination programmes, was suggested for greater reach and longevity; some interviewees said this had already been tried for HIV elimination. Interviewees had further suggestions for ***promoting hepatitis C testing to the public and patients***, including targeted campaigns with tailored messaging in practices and communities, and making other GPs aware of these campaigns. Some suggested that hepatitis C testing could be part of broader invitations for a health check / liver test, which might appeal more widely.

Finally, a need for alternative types of ***community clinics with GPs/pharmacists prescribing DAAs*** was suggested as likely to meet the needs of some patients better than conventional primary care practices.

> “I would like it that we could actually prescribe the drugs as well. We can’t at the moment, we can only do it through the ODN, but I think actually having you know some-a few GPs with a special interest potentially being able to prescribe the drugs for the patients, I think it would make it, I know they can deliver them and things, but it’s that engagement and getting them to then continue to take them and things which I think is really important.” (Participant 7).

Suggestions for special community GP or pharmacy clinics in areas of need cited potential benefits, including reducing the need for patients to travel; enabling GPs/pharmacists to establish trust with patients; and enabling GPs to maintain contact throughout treatment.

## Discussion

In this study we interviewed GPs with expertise in hepatitis C to understand the barriers and facilitators to identifying and (re)engaging patients with hepatitis C via primary care.

### Barriers and facilitators to hepatitis C care for all GPs

At a system level, similarly to previous research [13] the barriers interviewees cited included a lack of financial incentives for hepatitis C, workload pressures facing primary care, and difficulties applying patient searches that rely on complex medical terminology for hepatitis C. In addition, interviewees said that there are a lack of clear responsibilities or guidelines about how often to follow up patients living with or at risk of hepatitis C infection.

Provider-level barriers and facilitators likewise had some similar findings as those in previous studies [13, 22], indicating that they remain relevant. For example, GPs may still have insufficient training in hepatitis C, may not have capacity to prioritise hepatitis C over other duties, and face difficulty contacting and communicating with patients about hepatitis C. As also found in previous research [13, 22], facilitators included the holistic approach and local accessibility of GPs as a strength, and interviewees emphasised that successful patient (re)engagement relies on being able to understand patient needs and offer appointment flexibility.

Additional facilitators for identifying and (re)engaging patients included digital resources such as patient record system reminders, pop-ups, and automated blood testing panels. Interviewees said these digital solutions might address some of the capacity and capability barriers faced by GPs (e.g., forgetting to offer testing or not having time to read through patients’ notes in a routine appointment). This is consistent with other research demonstrating the advantages of combining holistic care with a digital care tool [36].

Interviewees’ suggestions for the future were to improve the functionality of digital tools to enable hepatitis C testing and referrals to become routine practice.

### Hepatitis C Champion GP roles

In addition to the above factors that impact all GPs, we identified some novel themes related to the hepatitis C GP Champions. Champions presented advantages and disadvantages to their roles in establishing collaborations and data-sharing with secondary care (ODNs), other GPs in Primary Care Networks, and local drug and alcohol services.

Champions said their unique roles are facilitating extra capacity in their schedules and enabling them to overcome some of the cited provider-level barriers. Due to the training, support, and experience they have gained, they expressed confidence in talking to patients about hepatitis C and feelings of professional reward (also found in a study where GPs were enabled new prescribing roles [37]).

However, they also reported novel barriers. GP Champions face challenges refining searches of patient records due to scale of worked needed across regions, and in determining effective and ethical patient communication strategies. Interviewees’ suggestions were to build on successes through further expansion of the GP Champion initiatives, and incentivisation, resources, and training for hepatitis C in primary care.

### Shared care models

Some GP interviewees reported on successfully sharing care with secondary care (ODNs). This meant either sharing efforts to identify and re-engage patients for referral to secondary care, or secondary care enabling GPs to prescribe treatment in primary care. Interviewees also emphasised success in the use of multi-disciplinary and collaborative outreach methods and community clinics for treatment, with suggestions there need to be more of these in areas of higher need. If GPs and pharmacists could prescribe DAAs and have more local and flexible clinics, interviewees suggested this would enable GPs to establish trust with patients by treating them holistically.

These findings about shared care align with existing guidance and research on how to engage in shared care [5, 28], and research showing the mutual benefits for patients and providers of doing so [22, 23, 37]. One benefit is that primary care providers have a special role, compared to secondary care, in helping the patient consider the broader health and wellbeing benefits from treating hepatitis C [22]. These suggestions coincide with the new UK Government 10-year plan for the NHS [38], and the HCV Action Collective report on the maintenance of elimination of hepatitis C [39], which emphasise joint initiatives for community and preventative health care.

### Strengths and limitations

By design, the participants in this study were experienced in hepatitis C management in primary care. This resulted in useful insights, and we gathered an understanding of the GP Champion initiative. However, interviewees also reflected on how their experiences may not be typical of other GPs. Finally, the findings were specific to England and the NHS, and may not all be applicable internationally; however, we have discussed some similarities and differences to international research.

## Conclusion

This study explored barriers and facilitators faced by GPs who are engaged in identifying and (re)engaging patients living with hepatitis C. Interviewees reported that their funding and the support from other GP Champions enabled them to overcome some of the barriers facing GPs, particularly capacity and knowledge limitations. Additionally, they reported professional benefits such as improved confidence in working with patients with hepatitis C and finding the work personally rewarding. They also reported new challenges and persistent challenges that have not yet been addressed. These included: difficulties coordinating efforts to find and (re)engage patients across regions; searching patient records and the complexity of digital reminders to (re)offer patients hepatitis C tests; and the need for a holistic and flexible approach to meet patient needs. Furthermore, their suggestions for future work towards hepatitis C elimination entailed the desire to see blood-borne virus elimination campaigns and services join up, further incentives and training for hepatitis C, and for more community multi-disciplinary clinics to provide holistic care, including DAA prescriptions. Therefore, GPs with funded capacity may help (re)engage patients in care, but system and provider-level barriers remain and need to be overcome; combined conventional and outreach approaches and joint efforts between services/sectors were suggested as means to overcome some of these barriers in addition to further funding for primary care.

## Declarations

### Ethics approval and consent to participate

The study was approved by the UKHSA Research Ethics and Governance Group (ID: 575). Participants were informed about the study and provided electronic written consent.

### Consent for publication

All participants consented to have anonymised quotes published.

### Availability of data and materials

Interview transcripts are stored by the corresponding author and can be made available upon reasonable request.

### Authors’ contributions

AS contributed to Conceptualization, Methodology, Formal analysis, Investigation, Data Curation, Writing - Original Draft, Writing - Review & Editing, Supervision, Project administration. CH contributed to Conceptualization, Methodology, Formal analysis, Writing - Review & Editing. DM contributed to Conceptualization, Methodology, Investigation, Data Curation, Writing - Review & Editing. CL contributed to Conceptualization, Methodology, Supervision, Writing - Review & Editing. RS, AN, and MD contributed to Conceptualization, Methodology, Writing - Review & Editing.

## Data Availability

All interview transcript files will be available from OSF (https://osf.io/dnwv8/overview).

## Acknowledgements

Thank you to all the GPs who participated in this study.

## List of Abbreviations

DAA: direct-acting antiviral
GP: general practitioner
hepatitis C: hepatitis C virus
ODN: operational delivery network

## Footnotes

1 The <u>PSI: Patient Search Identification tool</u> searches for Hepatitis C risk factors in electronic patient record systems.

